# Association between antinuclear antibodies (ANA) patterns and extractable nuclear antigens (ENA) in patients with autoimmune diseases

**DOI:** 10.1101/2024.01.04.24300829

**Authors:** Abdul Rehman Haris, Hamid Nawaz Tipu, Dawood Ahmed, Muhammad Abdullah Nabeel, Muhammad Shoaib Akhtar

## Abstract

**Background:** Antinuclear antibodies (ANA) and extractable nuclear antigens (ENA) are crucial biomarkers for the diagnosis of autoimmune diseases (AID) such as systemic lupus erythematosus (SLE), Sjogren’s syndrome, systemic sclerosis, and polymyositis. In the present study, we assessed the most frequent ANA patterns associated with the most detectable ENA antigen (Ag) that could be used as a diagnostic and efficient prognostic marker of AID.

**Materials and Methods:** The primary objective of this study was to investigate the association between immunofluorescence (IF) ANA and ENA in patients with AIDs. This was a retrospective cross-sectional study. The study was performed at the Immunology Department of the Armed Forces Institute of Pathology, Rawalpindi. Retrospective data from 76 patients were tested for ANA and ENA from June 2020 to Nov 2020.

**Results:** A total of 76 patients comprising 14 (18.4%) males and 62(81.6%) females were tested for AIDs. The most frequent pattern among AID patients was coarse speckled, followed by the peripheral ANA pattern. The most frequent ENA Ags were Sjogren’s syndrome A (SSA) and B (SSB). SSA was significantly associated with coarse speckled and peripheral ANA patterns, whereas SSB was associated with coarse speckled ANA patterns. These associations are relevant for accurate diagnosis of autoimmune diseases.

**Conclusion:** SSA was associated with coarse speckled and peripheral ANA patterns, whereas SSB was associated with coarse speckled ANA patterns. The ANA patterns were significantly associated with ENA antigens.

## Introduction

The primary objective of the immune system is to protect the body from various pathogens. The immune system recognizes foreign objects through pathogen recognition receptors (PRRs) such as Ag. In response to these Ag, the immune system produces specific antibodies. Normal adults usually exhibit tolerance to self-antigens. These antigens present to the cell during fetal life and are recognized as the “self”. In some situations, tolerance to these antigens may be lost, and immune reactions may start against these antigens. A key stage in all autoimmune disorders is the activation of CD4-positive helper T cells. If this occurs, these T cells develop into helper T cells, such as Th1, Th2, and other types, and produce inappropriate inflammation. Autoantibody-producing B cells are then activated (1).

The frequency of autoimmune diseases in the Pakistani population was 55% in different age groups (2). Similar studies in India and other neighboring countries have reported the prevalence of ANA to range from 5% to 38% in different geographic regions (3–6). On the other side, Eurasian countries also reported similar statistics (15% to 33%) (7–10). The prevalence of ANA-positivity reported in the United States was 13.8% (11). The increased frequency of autoimmune diseases shows that ANA is an important diagnostic test. However, the prevalence of ANA in healthy individuals, especially young people, has increased over the last decade. In healthy individuals, ANA occurs at low concentrations, which demands a cut-off value for ANA (12).

Ada Man and his colleagues developed an algorithm for autoimmune diseases which cancels the ENA and anti-ds DNA tests if ANA was negative in any sample. This algorithm allows the laboratory to save up to 30% of the cost of an individual (13).

Systematic autoimmune rheumatic disorders (SARDs) are heterogeneous disorders, including SS and SLE (14). These disorders can be diagnosed based on the presence of specific antibodies against self-antigens. These antibodies are commonly detected using ANA, anti-ENA, and anti-double-stranded DNA (ds-DNA) antibodies.

ANA is performed for specific antibodies against the cellular nucleus such as anti-ds-DNA antibodies, anti-SSA, autoantibodies to SSA (Ro), and anti-SSB. These antibodies are diagnostic and prognostic biomarkers for monitoring autoimmune diseases. ANA has been detected in human laryngeal carcinoma HEP-2 cells and rat liver cells (15). When ANA was positive, Anti ENA was performed to confirm the diagnosis. Ideally, the ANA is a critical biomarker for the diagnosis of SARD such as systemic sclerosis, SLE, mixed connective tissue disorder (MCTD) and SS (14). The ANA assay is used to detect specific autoantibodies against nucleus or nuclear components. In addition, some cytoplasmic antibodies, such as mitotic cellular apparatus antibodies, are also detected by ANA. (16). The five targets of the ANA test include histone proteins, ds-DNA, DNA/histone complexes (nucleosomes), various nuclear enzymes, and ribonucleoprotein (RNP) (17).

ANA show different staining patterns, such as homogenous, nucleolar, and speckled. Based on these staining patterns, ANA is subdivided into nucleolar (N-ANA), homogenous (H-ANA), speckled (S-ANA), peripheral (P-ANA), and centromere (C-ANA). The association between homogenous ANA and AID or normal cells is controversial (18). As ANA helps to determine the presence or absence of autoantibodies, ENA helps diagnose the exact AID. ENA profiling is important for detecting autoantibodies against proteins within the nucleus. Moreover, ENA profiling helps to analyze the prognosis of AID. (19)

There is no documented evidence of the prevalence of AID in Pakistan. Therefore, it is important to conduct further research on AID to determine their epidemiology. From this perspective, the present study reviewed the data of patients with AIDs reported at the Immunology Department of the Armed Forces Institute of Pathology in Rawalpindi, Pakistan. This study aimed to determine the association between indirect immunofluorescence ANA patterns and ENA frequencies in patients with AID.

### Methodology

This was a retrospective cross-sectional study. The study was conducted at the Immunology Department, Armed Forces Institute of Pathology (AFIP), Rawalpindi, a training institute affiliated with the Armed Forces Post Graduate Medical Institute (AFPGMI), Rawalpindi. All patients tested for both ANA and ENA at the AFIP were referred from the Combined Military Hospital (CMH) Rawalpindi and other military hospitals. The study duration was six months, from June 2020 to Nov 2020. All the samples tested for both ANA and ENA were included in this study.

### ANA Detection

The ANA testing protocol involved a previously reported in-house indirect immunofluorescence (IIF) assay (20). Slides coated with substrates containing rat liver and/or kidney cells were used. Plasma and serum samples were diluted in a buffer solution. The diluted samples were then applied to substrate-coated slides. Slides containing the sample-substrate mix were incubated at room temperature for 20 min in a moist chamber, followed by washing. The slides were then rinsed in phosphate-buffered saline (PBS) for 20 min at room temperature in the dark. Fluorescent-tagged, diluted, conjugated anti-human antibodies were dispensed on each slide. Slides were again incubated in a moist chamber at room temperature in the dark, followed by rinsing with PBS. The slides were mounted in 1:10 glycerol in PBS. The slides were then examined under a fluorescence microscope to determine the presence and pattern of ANA.

### ENA Profiling

ENA profiling included the detection of SSA (Ro), Ro-52, Ro-60, SSB, Scl-70, Jo-1, anti-RNP, and anti-Smith antibody (Sm) by western immunoblot assay. ENA testing was performed using a commercially available strip-based immunoblotting kit (EuroImmune cat # EA 1590-1208-1G) to ensure accurate and reliable results. The manufacturer-recommended protocol was followed for the ENA profiling of all samples. The ENA developed on the strips was compared with manufacturer-provided control bands. Quality control measures, including the use of positive and negative controls provided in the kit, were implemented to ensure reliability of the results.

### Data Entry and Statistical Analysis

Data were coded and entered into Microsoft Excel 2020 and Statistical Package for Social Sciences (SPSS Version 22). Fisher’s Exact test was used to detect the association between ANA and ENA and between different ANA patterns and ENA antigens separately. The ROC curve was drawn to determine the sensitivity and specificity of ENA.

## Results

A total of 76 patients (14 males and 62 females) were tested for ANA and ENA. Of these, 12 (85.7%) were ANA-positive among males, while 50 (80.6%) were positive among females. Six males (42.9%) and 40 females (64.6%) were ENA-positive. The frequency of ANA patterns observed in both sexes is shown in Table 1.

**Table 1.** Gender distribution of ANA Patterns.

| ANA Patterns |  |  |  |  |  |  |
| --- | --- | --- | --- | --- | --- | --- |
|  | Coarse speckled | Fine Speckled | Homogenous | Nucleolar | Centromere | Peripheral |
| Male | 5 | 1 | 1 | 1 | 0 | 3 |
| Female | 24 | 4 | 4 | 7 | 3 | 8 |
| Total | 29 | 5 | 5 | 8 | 3 | 11 |

Coarse speckles were the most prevalent pattern among the males and females. The least frequent pattern in both sexes was the anti-centromere pattern (CEN-ANA).

The percentage of ENA antigens among male and females are given in figure 1.

**Figure 1:**
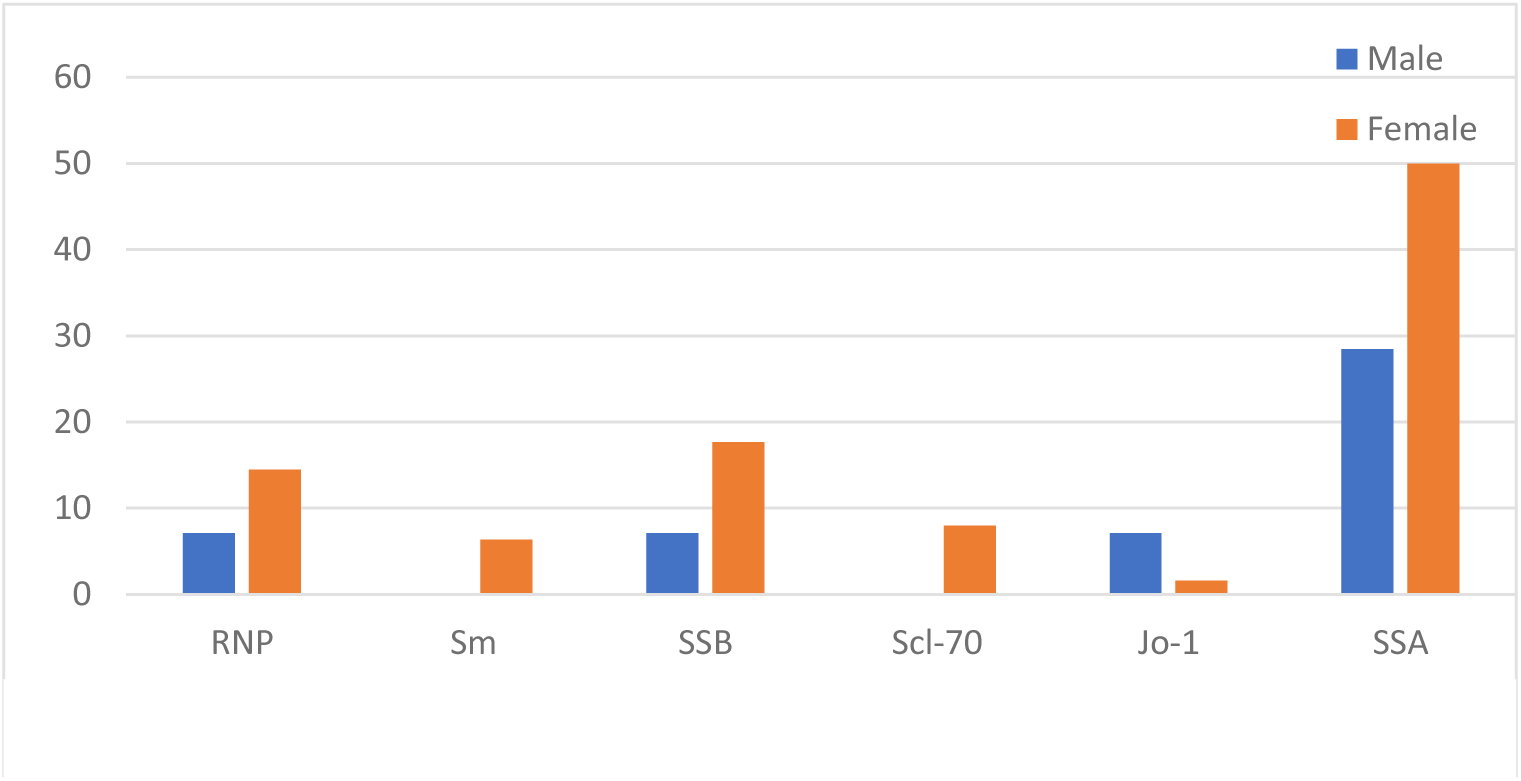
Gender Distribution of ENA Antigens.

**Figure 2:**
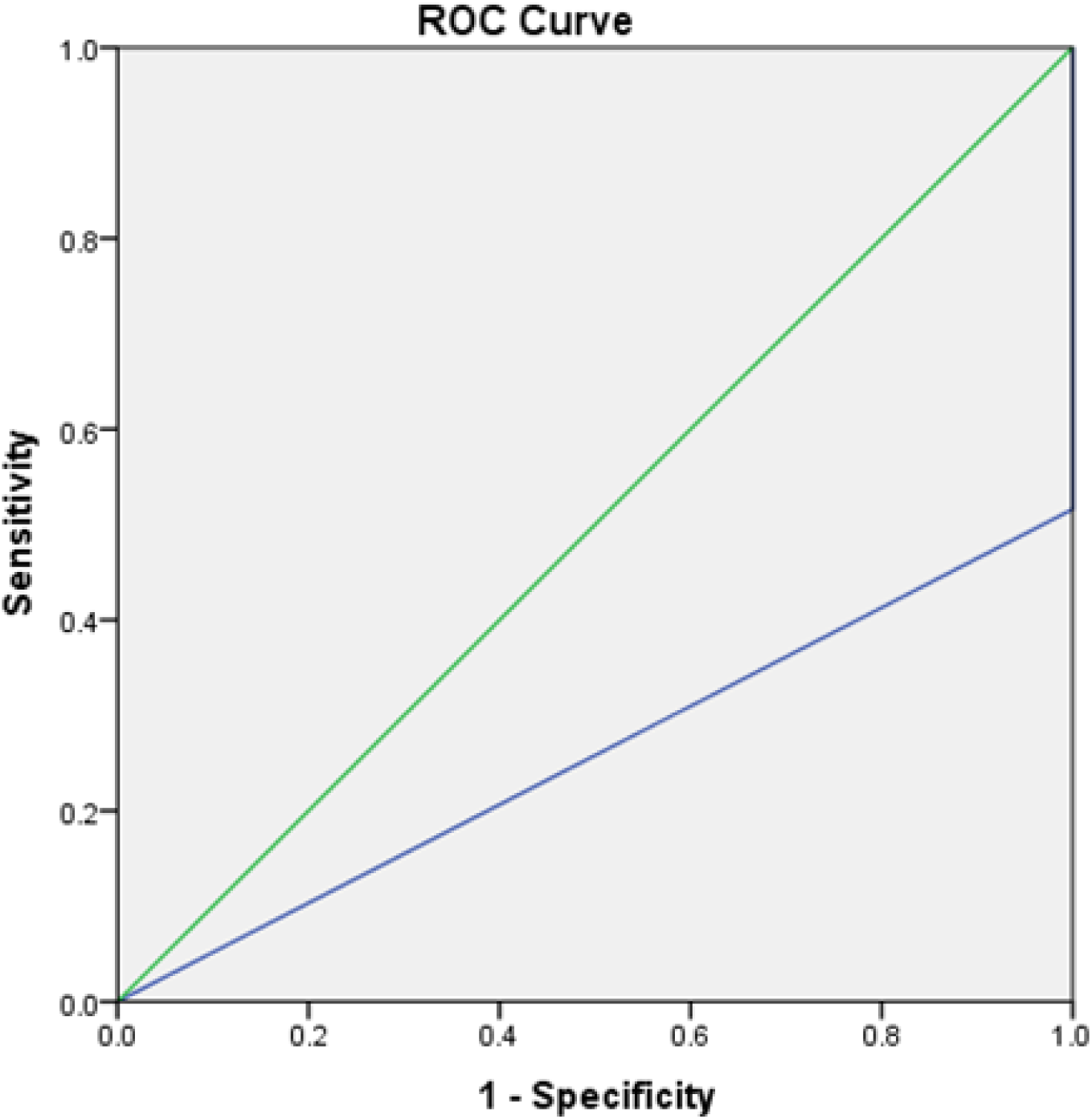
Sensitivity and Specificity of ENA.

Furthermore, to determine the most prevalent ENA among different ANA patterns, we carried out a study of ENA-detecting Ags, including SSA, SSB, Scl-70, Jo-1, RNP, and Sm. The most prevalent Ag among both sexes was SSA. The least frequent Ag was Sm and Scl-70 in males and J0-1 in females.

ENA was detectable at different percentages in different patterns. The prevalence of these Ags in the different patterns is shown in Table 2. The most prevalent pattern among ANA was coarse speckled, while the highest prevalence of ANA patterns in ENA Ag was peripheral ANA in SSA Ag of ENA.

**Table 2:** Prevalence of ENA Antigens and ANA Patterns.

|  |  | Coarse<br>Speckled | Fine<br>Speckled | Homogenous | Nucleolar | Centromere | Peripheral | Negative |
| --- | --- | --- | --- | --- | --- | --- | --- | --- |
| RNP | Negative | 23 | 5 | 4 | 8 | 3 | 9 | 14 |
|  | Positive | 6 | 0 | 1 | 0 | 0 | 2 | 1 |
| Sm | Negative | 29 | 5 | 3 | 8 | 3 | 9 | 15 |
|  | Positive | 0 | 0 | 2 | 0 | 0 | 2 | 0 |
| SSB | Negative | 29 | 4 | 4 | 7 | 3 | 7 | 10 |
|  | Positive | 0 | 1 | 1 | 1 | 0 | 4 | 5 |
| Scl-70 | Negative | 26 | 4 | 5 | 7 | 3 | 11 | 15 |
|  | Positive | 3 | 1 | 0 | 1 | 0 | 0 | 0 |
| Jo-1 | Negative | 29 | 5 | 5 | 8 | 3 | 11 | 13 |
|  | Positive | 0 | 0 | 0 | 0 | 0 | 0 | 2 |
| SSA | Negative | 22 | 3 | 2 | 6 | 3 | 2 | 3 |
|  | Positive | 7 | 2 | 3 | 2 | 0 | 9 | 12 |

### Sensitivity of ENA

The estimated sensitivity and specificity of the ENA test for SLE are 100% and 86%, respectively. The sensitivity and specificity for other rheumatic diseases were 42% and 85%, respectively (21). According to the kit protocol for total ENA, the ENA kit sensitivity was 91%, and specificity was >99%. The ROC curve showed that there was 51.6% concordance between ENA and ANA, and the area under the curve (AUC) was 0.258. The calculated AUC shows that sensitivity or specificity is not significant for the following data, and there is a chance of false-positive or false-negative results of ENA.

### Relationship between ANA and ENA

Fisher’s Exact Test was performed to evaluate the relationship between ANA patterns and ENA. A significant relationship between the ENA and ANA patterns was observed (p=0.0069). Furthermore, ANA patterns were individually observed in association with ENA Ag. Antigen SSA was associated with two ANA patterns: coarse speckled and peripheral. SSA subtypes were also associated with ANA patterns; SSA (R0) was associated with coarse speckled and peripheral patterns, and SSA (Ro-52) was associated with peripheral patterns. SSB(La) was associated with coarse speckled and Sm was associated with a homogenous pattern. The p-values of all the calculations are given in Tables 3 and 4. In contrast, 26.3 % of ANA-negative patients were ENA-positive.

**Table 3:** P-values of ANA Patterns with ENA Antigens.

|  | Coarse<br>Speckled | Fine<br>Speckled | Homogenous | Nucleolar | Centromere | Peripheral |
| --- | --- | --- | --- | --- | --- | --- |
| **RNP** | 0.16 | 1 | 0.51 | 0.48 | 1 | 0.6 |
| **Sm** | 0.2 | 1 | **0.02** | 0.6 | 1 | 0.09 |
| **SSA** | **0.004** | 1 | 0.65 | 0.9 | 0.2 | **0.019** |
| **SSB** | **0.002** | 1 | 1 | 0.5 | 1 | 0.06 |
| **Scl-70** | 0.36 | 0.2 | 1 | 0.8 | 1 | 1 |
| **Jo-1** | 0.52 | 1 | 1 | 0.7 | 1 | 1 |

**Table 4:** P-values of ANA Patterns with SSA Subtypes.

|  | SSA (Ro) | SSA (Ro-52) | SSA(Ro-60) |
| --- | --- | --- | --- |
| **Coarse speckled** | **0.006** | 0.5 | 0.7 |
| **Fine speckled** | 1 | 1 | 1 |
| **Homogenous** | 0.33 | 1 | 1 |
| **Nucleolar** | 0.7 | 0.5 | 0.5 |
| **Centromere** | 0.5 | 1 | 1 |
| **Peripheral** | **0.006** | **0.04** | 1 |

## Discussion

In this study, we found that coarse speckled and peripheral patterns were the most frequent ANA patterns among AID patients. The frequency of ENA in all ANA patterns was significant at p <0.05.

The presence of ANA in the blood constitutes a significant criterion for the diagnosis of AID. The identification of the ANA subtype or ENA Ag plays a key role in the diagnosis of specific CTD. The present study verified the profiles of patient samples tested for ENA antibodies and correlated them with ANA patterns. The most common patterns were coarse speckled and peripheral ANA patterns (38.2% and 14.5 %, respectively). Among ENA, the most prevalent Ag was SSA, which was associated with coarse speckled and peripheral patterns of ANA (19). Another study also reported that coarse speckled ANA patterns are associated with SSA antibodies (22). The second most prevalent ENA antigen was SSB, which was associated with a coarse speckled pattern. Fisher’s exact test of ENA and ANA was found to be significant at p < 0.05. In contrast, 26.3 % of ANA-negative patients were ENA-positive. This conclusion was also supported by Li (23). It is estimated that some individuals who are ANA negative may be positive for anti-cardiolipin, or anti double stranded DNA.(24).

A comprehensive understanding of the significance of various patterns can assist clinicians in confirming the diagnosis of AID. Notably, a homogeneous ANA pattern has been observed in association with SLE (25). These patterns mainly result from autoantibodies against the nucleus or the nuclear material (26). Speckled ANA has been reported to be the dominant immunofluorescence ANA pattern in SLE patients (19). The most frequent pattern identified in this study was speckled; therefore, the results were consistent with this study. Similarly, the highest prevalence of ENA Ag was observed in SSA, which was also reported by (27).

The determination of ENA contributes to improved differentiation among different types of autoimmune rheumatic diseases (ARD). ANA positivity with dsDNA or Sm positivity is a diagnostic criterion for SLE. Similarly, the presence of RNP autoantibodies is an important marker for MCTD diagnosis. In addition, anti-SSA and anti-SSB antibodies are significant immune markers for the diagnosis of Sjögren’s syndrome, subacute cutaneous SLE, and neonatal lupus syndrome. The Jo-1 autoantibody against histidyls RNA synthetase is capable of acting as an immunomarker linked with polymyositis or dermatomyositis, and Scl-70 facilitates the diagnosis of systemic sclerosis (28).

Fisher’s exact test was used to determine the association between ENA antibodies and ANA patterns. There was no significant association between RNP and ANA patterns. The p-values for coarse speckled, fine speckled, homogenous, nucleolar, centromere, and peripheral were 0.16, 1, 0.51, 0.48, 1, and 0.6, respectively.

A significant association was found between the anti-Smith antibody and Homogenous ANA pattern with a P-value 0.02 (less than 0.05). However, there was no significant association between anti-Smith antibody and coarse speckled, fine speckled, nucleolar, centromere, and peripheral, with P-values of 0.2, 1, 0.6,1, 0.09, respectively.

There was also a significant association between anti-SSA and ANA coarse speckled and peripheral patterns with p-value 0.04 and 0.019 respectively. However, the fine speckled, homogenous, nucleolar, and centromere patterns were not associated with SSA, with P-values of 1. 0.65, 0.9, and 0.2, respectively). SSA subtypes were also checked for association and it was found that SSA(Ro) is associated with coarse speckled ANA pattern with p-value 0.006, and SSA (Ro) and SSA (Ro-52) have significant associations with ANA peripheral patterns with p-values 0.006 and 0.04, respectively.

SSB and ANA coarse speckled patterns had a significant association, with a p-value of 0.002 (less than 0.05), while SSB and ANA fine speckled, homogenous, nucleolar, centromere, and peripheral patterns had no significant relationship, with p-values of 1, 1, 0.5, 1, and 0.06, respectively. Scl-70 and J0-1 did not show any significant relationship with ANA patterns.

Interpreting the research results based on a small dataset can present certain limitations and uncertainties. Although the initial findings may provide valuable insights, it is important to acknowledge the potential for variations and changes when conducting the same research on a larger and more diverse dataset. With a larger sample size, it is possible to capture a wider range of perspectives and factors that can influence the overall outcomes and generalizability of the research. Therefore, expanding the research to include a larger dataset would enhance the reliability and validity of the findings, providing a more comprehensive understanding of the phenomenon under investigation.

The findings of my research indicate that ANA can also function as a diagnostic test for specific antibodies and has significant implications for clinicians. First, by establishing the diagnostic accuracy of ANA, clinicians can use this test as a valuable tool for their diagnostic arsenal. Second, it offers a more cost-effective and accessible approach than relying solely on ENA.

## Conclusion

The most frequent ANA pattern was the coarse speckled peripheral pattern. The most prevalent Ag among both sexes was SSA. The least frequent Ag was Sm and Scl-70 among males and J0-1 among females. The most prevalent ENA antigens were SSA and SSB antigens. SSA was associated with coarse speckled and peripheral ANA patterns, whereas SSB was associated with coarse speckled ANA patterns. The ANA patterns were significantly associated with ENA antigens.

Interpreting research results from a small dataset has limitations. Conducting the same research on a larger, diverse dataset can capture a wider range of factors, improving outcomes and generalizability. Expanding research with a larger dataset enhances reliability and provides a comprehensive understanding of this phenomenon.

## Data Availability

All data produced in the present study are available upon reasonable request to the authors

## References

1. Warren Levinson PC-H, Elizabeth A. Joyce, Jesse Nussbaum, Brian Schwartz. Review of Medical Microbiology and Immunology: McGraw-Hill Medical; 2018.

2. Imran K, Loya A, Hameed M, Siddiqui IA, Sheikh UN, Sheikh U. The Frequency of Immunofluorescence Antinuclear Antibody Patterns and Extractable Nuclear Antigen: Experience From a Large Laboratory in Pakistan. Cureus. 2023;15(1).

3. Prapinjumrune C, Prucktrakul C, Sooktonglarng T, Thongprasom K. Serum antinuclear antibody in adult Thais. Gerodontology. 2017;34(1):86–9.

4. Sebastian W, Roy A, Kini U, Mullick S. Correlation of antinuclear antibody immunofluorescence patterns with immune profile using line immunoassay in the Indian scenario. Indian Journal of Pathology and Microbiology. 2010;53(3):427.

5. Guo Y-P, Wang C-G, Liu X, Huang Y-Q, Guo D-L, Jing X-Z, et al. The prevalence of antinuclear antibodies in the general population of china: a cross-sectional study. Current Therapeutic Research. 2014;76:116–9.

6. Minz RW, Kumar Y, Anand S, Singh S, Bamberi P, Verma S, Shegal S. Antinuclear antibody positive autoimmune disorders in North India: an appraisal. Rheumatology international. 2012;32:2883–8.

7. Mengeloglu Z, Tas T, Kocoglu E, Aktas G, Karabörk S. Determination of anti-nuclear antibody pattern distribution and clinical relationship. Pakistan journal of medical sciences. 2014;30(2):380.

8. Sener AG, Afsar İ, Demirci M. Evaluation of antinuclear antibodies by indirect immunofluorescence and line immunoassay methods′: four years′ data from Turkey. APMIS. 2014;122(12):1167–70.

9. Peene I, Meheus L, Veys E, De Keyser F. Detection and identification of antinuclear antibodies (ANA) in a large and consecutive cohort of serum samples referred for ANA testing. Annals of the rheumatic diseases. 2001;60(12):1131–6.

10. Akmatov MK, Röber N, Ahrens W, Flesch-Janys D, Fricke J, Greiser H, et al. Anti-nuclear autoantibodies in the general German population: prevalence and lack of association with selected cardiovascular and metabolic disorders—findings of a multicenter population-based study. Arthritis Research & Therapy. 2017;19:1–9.

11. Satoh M, Chan EK, Ho LA, Rose KM, Parks CG, Cohn RD, et al. Prevalence and sociodemographic correlates of antinuclear antibodies in the United States. Arthritis & Rheumatism. 2012;64(7):2319–27.

12. Pashnina IA, Krivolapova IM, Fedotkina TV, Ryabkova VA, Chereshneva MV, Churilov LP, Chereshnev VA. Antinuclear autoantibodies in health: autoimmunity is not a synonym of autoimmune disease. Antibodies. 2021;10(1):9.

13. Man A, Shojania K, Phoon C, Pal J, de Badyn MH, Pi D, Lacaille D. An evaluation of autoimmune antibody testing patterns in a Canadian health region and an evaluation of a laboratory algorithm aimed at reducing unnecessary testing. Clinical rheumatology. 2013;32:601–8.

14. Stinton LM, Fritzler MJ. A clinical approach to autoantibody testing in systemic autoimmune rheumatic disorders. Autoimmunity reviews. 2007;7(1):77–84.

15. Kumar Y, Bhatia A, Minz RW. Antinuclear antibodies and their detection methods in diagnosis of connective tissue diseases: a journey revisited. Diagnostic pathology. 2009;4(1):1.

16. Andrade LEC, Damoiseaux J, Vergani D, Fritzler MJ. Antinuclear antibodies (ANA) as a criterion for classification and diagnosis of systemic autoimmune diseases. Journal of Translational Autoimmunity. 2022:100145.

17. Hu Z-D, Deng A-M. Autoantibodies in pre-clinical autoimmune disease. Clinica Chimica Acta. 2014;437:14–8.

18. Von Mühlen C, Tan E. Autoantibodies in the diagnosis of systemic lupus erythematosus. Semin Arthritis Rheum. 1995;5:323–58.

19. Alsubki R, Tabassum H, Alfawaz H, Alaqil R, Aljaser F, Ansar S, Al Jurayyan A. Association between antinuclear antibodies (ANA) patterns and extractable nuclear antigens (ENA) in HEp-2 cells in patients with autoimmune diseases in Riyadh, Saudi Arabia. Intractable & Rare Diseases Research. 2020;9(2):89–94.

20. Masood anwar Ma, waqar Farooq, ahmad khan, Waheed uz zaman, tariq Suhaib, ahmed Sajid, mushtaq Tahir, aziz ahmad, Sajjad hussain mirza, Mirza muhammad dawood, editor. Manual of Laboratory Medicine. Armed Forces Institute of Pathology rawalpindi 2005.

21. Slater CA, Davis RB, Shmerling RH. Antinuclear antibody testing: a study of clinical utility. Archives of Internal Medicine. 1996;156(13):1421–5.

22. Anis S, Fatima A, Jabbar SA, Arain T. Anti-ENA Antibodies, ANA Patterns, Anti-ds DNA results, and Clinical Diagnosis: A Laboratory and Clinical Audit. 2022.

23. Yeo AL, Ojaimi S, Le S, Leech M, Morand E. Frequency and clinical utility of antibodies to extractable nuclear antigen in the setting of a negative antinuclear antibody test. Arthritis care & research. 2022.

24. Rodríguez-Orozco AR, Bejar-Lozano C, Cortés-Rojo C. Antibodies to extractable nuclear antigens are detectable in a considerable number of sera that test negative for antinuclear antibodies. Archives of Pathology & Laboratory Medicine. 2022;146(2):143–4.

25. Sjöwall C, Sturm M, Dahle C, Bengtsson AA, Jönsen A, Sturfelt G, Skogh T. Abnormal antinuclear antibody titers are less common than generally assumed in established cases of systemic lupus erythematosus. The Journal of rheumatology. 2008;35(10):1994–2000.

26. Frodlund M, Dahlström Ö, Kastbom A, Skogh T, Sjöwall C. Associations between antinuclear antibody staining patterns and clinical features of systemic lupus erythematosus: analysis of a regional Swedish register. BMJ open. 2013;3(10).

27. Lora PS, Laurino CCFC, Freitas AEd, Brenol JCT, Montecielo O, Xavier RM. Antinuclear antibodies (ANA) immunofluorescent pattern s in HEp-2 cells on samples positive for anti-SSA/Ro. Revista Brasileira de Reumatologia. 2007;47(1):4–9.

28. Thompson D, Juby A, Davis P. The clinical significance of autoantibody profiles in patients with systemic lupus erythematosus. Lupus. 1993;2(1)yy:15–9.

